# DESIGN, DEVELOPMENT, AND VALIDATION OF A NANOBIOSENSOR TO DETECT CIRCULATING MICROBIOME FOR CARDIOVASCULAR DISEASE RISK ASSESSMENT

**DOI:** 10.1101/2023.08.05.23293588

**Authors:** Nazim Nazeer, Rakhi Dewangan, Kaniz Zaidi, Vikas Gurjar, Rajnarayan Tiwari, Pradyumna Kumar Mishra

**Author notes:** **Author for Correspondence: Prof. (Dr.) Pradyumna Kumar Mishra**, *MSc, PhD, Fulbright-Nehru Fellow (USA), AIMF (DBT-Wellcome Trust & AAS), FAMS*, **Scientist - F & Head**, Division of Environmental Biotechnology, Genetics & Molecular Biology (EBGMB), ICMR-National Institute for Research in Environmental Health (NIREH), NIREH Campus, Bypass Road, Bhauri, Bhopal - 462030 (MP), India.

## Abstract

Cardiovascular disease (CVD) is a serious worldwide health concern that necessitates the development of novel diagnostic techniques for early identification and personalized healthcare management. Even before the insights provided by gut microbiota, current research has demonstrated the importance of circulating microbiome (CMB) in the evolution of cardiometabolic illness risk and progression. We developed a nanobiosensor that uses specific labeled capture probes with perovskite quantum dots (PQDs) to detect the targeted 16S rRNA sequences in the peripheral milieu. With ideal applicability, specificity, and sensitivity, this sensor delivers unique insights into the presence and characterization of circulating microbiota signatures. Developing a nanophotonic microbiome detection method in body fluids may pave the way for creating a distinctive tool for CVD risk prediction for population-based screening programs in low and middle-income countries.

## INTRODUCTION

The microbiome (MB) is a vast and complex ecosystem of bacteria hypothesized to have a key role in various human pathophysiology (Srivastava et al., 2021). Previously, research focused on microbial communities in specific bodily locations such as the skin, oral cavity, and gut (Mishra et al., 2011; Sumida et al., 2022; Ullah Goraya et al., 2023). New research, however, has shed light on another important aspect known as the “circulating microbiome (CMB),” which contains a diverse array of bacteria, viruses, fungi, and other microbial entities, including bacterial DNA found in the bloodstream and other physiological fluids entered through the oral, intestinal mucosa, lungs, vaginal, and different routes, among others (Khan et al., 2022; Sciarra et al., 2023). The invasion of microbial signatures into the peripheral milieu has been linked to various illnesses and invasive medical procedures. For example, an imbalance or change in the composition of gut microbiota (dysbiosis) has been linked to the release of MB in the circulating milieu (Masenga et al., 2022). CMB and cell-free circulating DNA is thought to play a role in the etiology of various cardiovascular illnesses, including pericarditis, endocarditis, rheumatic pericarditis, hypertension, and atherosclerosis (Mishra and Kaur, 2023). One of the major processes by which the CMB affects cardiovascular health is the creation of metabolites such as trimethylamine (TMA), which is further oxidized in the liver by the enzyme flavin monooxygenase 3 to generate trimethylamine-N-oxide (TMAO) (Zhao & Wang, 2020). TMAO at high levels influences lipid metabolism, inflammation, and the function of endothelial cells lining blood vessels, causing atherosclerosis (Lawrence et al., 2022) (Figure 1). Furthermore, dysbiosis can cause metabolic changes that cause certain gut bacteria to create chemicals such as lipopolysaccharides, which can enter the circulation and trigger a systemic inflammatory response. Chronic inflammation is dangerous because it damages blood vessels, promotes plaque formation, and contributes to the development of CVDs (Velmurugan et al., 2020; Mishra et al., 2022a). Changes in microbial composition or community-derived components, such as circulating nucleic acids of xenogeneic origin, can be directly or indirectly associated with increased disease vulnerability. Pathophysiological consequences linked to CVDs may arise because of exogenous genetic material activating immunological signaling pathways. It is important to acknowledge that various factors, including genetics, sedentary lifestyle, diet, and environmental influences, influence CMB’s impact on disease development. These factors can affect the composition and dynamics of the CMB and can be identified using different methods (Hasan and Yang, 2019).

**Figure 1:**
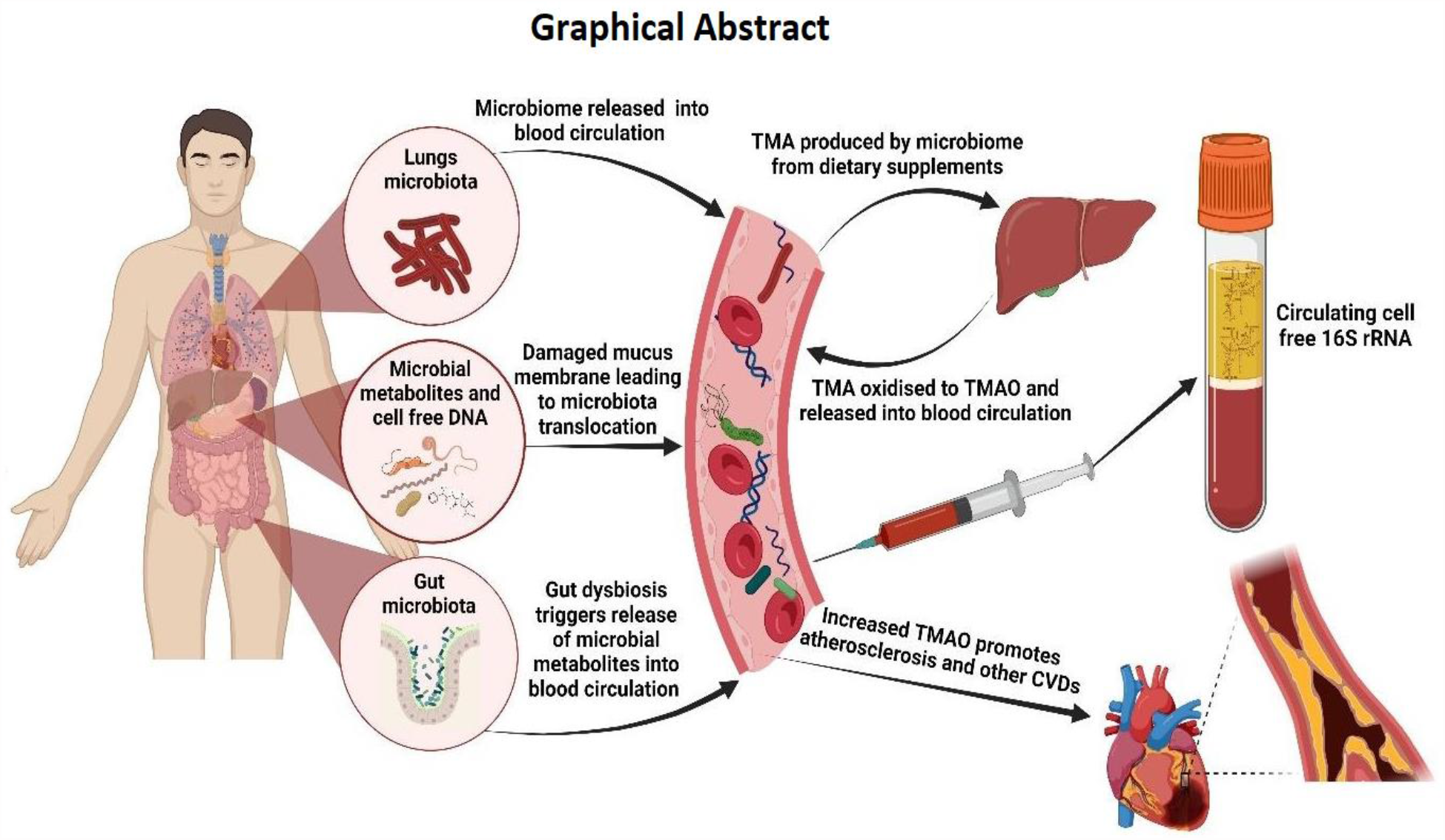
Circulating microbiome and possible patho-physiological implications in CVDs.

Researchers used conventional techniques such as nucleic acid amplification technology and sequencing to detect microbial communities and associate them with other metabolites in circulation for thorough characterization (Kabay et al., 2022). These processes usually entail circulating nucleic acid extraction, amplifying specific microbial genes or regions, and then employing sequencing methods (Lee et al., 2022). However, these methods are time-consuming and necessitate using bioinformatics-based skill sets, which may be challenging in a resource-constrained clinical setting (Ratre et al., 2022; Ratre et al., 2023). It is important to consider the limitations of these sequencing-based approaches when it comes to predicting risk for population-based screening programs in low and middle-income countries. It is now evident that sequencing-based testing may not always be able to provide straight-forward answers, especially when it comes to diseases that are linked to the microbiome. To fully understand these complex conditions, it is necessary to understand the integrated genomic-epigenomic-metabolomic mechanisms involved (Hasan and Yang, 2019; Mishra and Shandilya, 2020).

We herein report a novel fluorescence-based optical sensing approach that employs perovskite quantum dots (PQDs) as a component for the effective screening of MBs in circulation, and it shows tremendous potential by providing quick, cost-effective, and non-invasive microbiome profiling. PQDs have outstanding sensitivity and signal-to-noise ratios due to their unique photophysical features, such as strong fluorescence quantum yield and tunable emission wavelengths. Furthermore, the biocompatibility and ease of functionalization of PQDs increase their practical value. Our method employs streptavidin-conjugated PQDs that attach to biotinylated probes, resulting in a hybrid detection system that selectively targets the 16S rRNA for higher specificity in the detection of CMB.

## MATERIALS AND METHODS

### Reagents and solutions

The reagents and solutions used in this study were of the highest available grade. The PQDs were purchased from Merck Sigma-Aldrich, Inc. in St. Louis, MO, USA. EDC (1-ethyl-3-[3-dimethylaminopropyl] carbodiimide hydrochloride) and NHS (N-hydroxysuccinimide) were procured from Thermofisher Scientific in Waltham, Massachusetts, USA. The Pierce™ RNA 3’ End Biotinylation Kit was purchased from Thermo Fisher Scientific in Waltham, MA, USA. The polymerase chain reaction (PCR) primers, specifically the forward primer 5′-ATGGCTGTCGTCAGCT-3′ and the reverse primer 5′-ACGGGCGGTGTGTAG-3′, which targeted the long fragment of 16S rRNA, were obtained from Integrated DNA Technologies, Coralville, Iowa, USA and Eurofins Genomics, Anzinger Strasse,Germany. QIAMP circulating nucleic acid isolation kit was purchased from QIAGEN, Hilden, Germany. Master mix and SYBR Safe were purchased from Thermo Fisher Scientific (Waltham, MA, USA). 6X gel loading dye was obtained from Himedia Laboratory (Mumbai, MH, India). PrimeScript 1^st^ strand cDNA synthesis kit was purchased from Takara Bio Inc. (Shiga Japan).

### Structural characterization of PQDs

Prior to the nanohybrid preparation, the PQDs were characterized for particle/aggregate size and zeta potential analysis using the Malvern Panalytical Zetasizer Nano ZS with cuvette measurement cells (Malvern, United Kingdom). All measurements were conducted using vehicle mixtures.

### Circulating cell-free 16S rRNA characterization

The cell-free circulating RNA (ccf-RNA) was isolated from 10 blood samples using QIAMP circulating nucleic acid isolation kit following the manufacturer protocol. The study collected basic clinical data from all the volunteers aged 40 years and above, including information on hypertension, diabetes mellitus, and dyslipidemia. Venous blood samples were collected using standard venipuncture collection method and plasma was used for downstream processing. All laboratory procedures were performed as per the institutional ethical committee guidelines. The ccf-RNA was then subjected to quantification and subsequently cDNA preparation was performed using PrimeScript 1^st^ strand cDNA synthesis kit. The 16S rRNA strand was then amplified specifically using the appropriate primer (forward-ATGGCTGTCGTCAGCT, reverse-ACGGGCGGTGTGTAG (Integrated DNA Technologies, Coralville, Iowa, USA) from the prepared cDNAs. The master mix was used for the amplification of universal bacterial genes (16S rRNA) from the CMB by using Qiagen Rotor-Gene Q RT-PCR with the specific annealing temperature as reported elsewhere (Shandilya et al., 2022a).

### Preparation of biotinylated oligonucleotide capture probes

The target 16s rRNA complementary oligonucleotide probes were procured and biotinylated at 3′-position using Pierce™ RNA 3’ end biotinylation. Briefly, 10 pmol of oligonucleotide probes were incubated at 85 °C for 5 minutes, followed by immediate cooling. The incubated probes were then mixed with suggested volumes of biotinylated cytidine (Bis) phosphate, 30% PEG, 10X-RNA ligation buffer RNase inhibitor, and T4-RNA Ligase enzyme and were incubated at 4 °C overnight. The nuclease-free water (NFW) was added to facilitate the removal of RNA ligase subsequently with chloroform/isoamyl alcohol (24:1). After vortexing and high-speed centrifugation, the obtained aqueous phase was mixed with icy ethanol (100%), 5 M NaCl, and glycogen, and further incubated for 1 hour at -20 °C to precipitate the biotinylated probes. After separating the precipitated pellet, washing with 70% ethanol was performed and allowed to air-dry (Shandilya et al., 2022b). Finally, the pellets were resuspended in NFW and subsequently stored at -80 °C.

### Functionalization of PQDs and preparation of the nanohybrid

Before preparing the nanohybrid, carbodiimide chemistry was employed for the PQDs activation and reaction with streptavidin. For this EDC (1mg/mL) and NHS (1mg/mL) was mixed with toluene solution PQDs (10 mg/mL) in 1X-PBS and incubated for 1 hour at RT (room temperature). The activated PQDs were further treated with streptavidin and mixture was again-incubated for an additional 20 min at RT. This streptavidin-PQD conjugate mixture was treated with another aliquot prepared by mixing EDC and NHS, this resulted in optimized amine-carboxyl coupling conjugation reaction efficiency. The mixture was further stirred at RT for 1 hour. After completion of the reaction, excess PQDs were removed from streptavidin-coated PQD conjugates via centrifugation. Removed the supernatant to get streptavidin-coated PQD pellets and was stored at 4 °C for further utilization (Shandilya et al., 2021a). After the immobilization of streptavidin to the PQDs surface, pre-biotinylated oligonucleotide probes conjugation with streptavidin-biotin using coupling chemistry was performed. This step signifies analyzing the targeted PQDs i.e., 16S rRNA in every sample. Briefly, streptavidin-tethered PQDs were mixed with PBS having pH 7.4 using vortex mixing to achieve a thorough and uniform dispersion. Then the biotinylated probe was introduced into the mixture at a ratio of 1:2 (streptavidin-tethered PQDs to biotinylated probes). Streptavidin and biotin allow for strong and specific binding interaction. This mixture was subjected to vortexing for 1 hour at RT to facilitate efficient binding between the streptavidin and biotinylated probes and further incubated overnight at 4 °C allowing ample time for the streptavidin-biotin coupling to occur to provide the nanohybrid structure (Shandilya et al., 2020). This nanohybrid was capable of effectively detecting the CMB (16S rRNA) in the samples (Figure 2).

**Figure 2:**
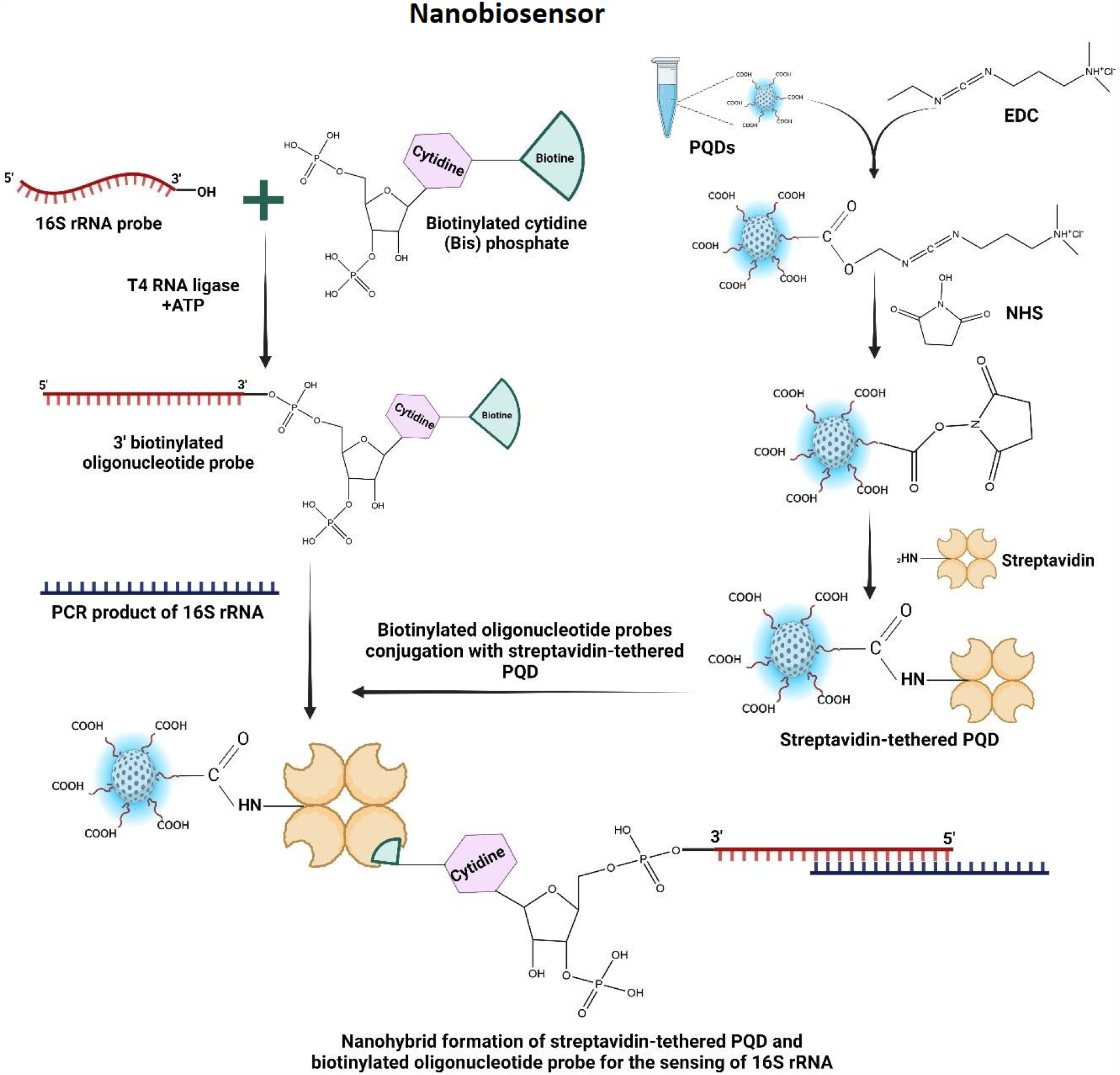
Sensing of 16S rRNA with the nanohybrid using strep-PQD linked with biotin-oligonucleotide probe.

### Co-localization analysis

The capability of the developed fluorescence-based optical sensing and specific identification of 16s rRNA among the circulating CMB was assessed by fluorescence microscopy and determined using fluorometry. For microscopy analysis, streptavidin-tethered PQDs biotinylated oligonucleotide nanohybrid was mixed with the 16S rRNA targets using Olympus high-resolution fluorescent microscope (Shinjuku City, Tokyo, Japan). This nanohybrid served as a probe with specificity for the target PCR product, allowing for the visualization of the specific 16S rRNA sequence. To enhance the signals for specific detection, propidium iodide was added to the mixture as an intercalating dye. Frost-free glass slides from Corning were used for imaging. To protect the sample and maintain optimal conditions, a cover slip was gently placed over the mixture on the glass slide. With the prepared slides in place, fluorescence microscopy analysis was conducted using appropriate filters (Mishra et al., 2022b).

### Applicability analysis

For the applicability experiments, Tecan Spark multimode reader (Männedorf Switzerland) was used. The first well was utilized as a blank, and 100 μl of 1X PBS was added for background reference (Shandilya et al., 2021b). In the subsequent wells, different combinations were prepared to evaluate the efficiency of streptavidin-conjugated PQDs in specific 16S rRNA detection. In the second well, a mixture of 90 μl of 1X PBS and 10 μl of PQDs was added as a control. The third well contained 90 μl of 1X PBS, followed by 10 μl of streptavidin-conjugated PQDs, with careful mixing using a pipette to avoid well wall contact. To examine the probe-target complex, the fourth well was loaded with 86 μl of 1X PBS, 10 μl of streptavidin-conjugated PQDs with a biotinylated probe, and 2 μl of the PCR product. The contents were thoroughly mixed before adding the propidium iodide dye. After a 10- to 15-minute incubation period, the microplate was placed in a multimode reader for data collection. The reader facilitated fluorescence analysis, allowing the assessment of specific 16S rRNA detection and interaction with the streptavidin-conjugated PQDs and the intercalating dye (PI). This experimental method provided important information about how streptavidin-conjugated PQDs could be used and how well they worked in detecting 16S rRNA.

### Sensitivity analysis

For the sensitivity test, a series of dilutions of the PCR product was prepared in a tube, ranging from 1000 ng to 0.1 fg. The purpose was to determine the detection limit of streptavidin-conjugated PQDs with biotinylated probes for 16S rRNA. The microplate was set up as follows: the first well served as a blank with 100 μl 1X PBS for background measurements. In the second well, 86 μl 1X PBS was added, followed by 10 μl streptavidin-conjugated PQDs with a biotinylated probe. Subsequently, 2 μl PCR products with varying concentrations (ranging from 1000 ng to 0.1 fg) were added to different wells, along with 2 μl PI dye. After incubating the plate for 10–15 minutes, it was placed in the multimode reader for data collection. The reader allowed for sensitive fluorescence analysis, enabling detection limit determination of streptavidin-conjugated PQDs with biotinylated probes. Following the experiment, a data analysis, visualization, and processing method was established in the Spark Control Magellan software (Shandilya et al., 2022a). This sensitivity experiment provided important insights into the performance and detection capabilities of the nanohybrid system, shedding light on its effectiveness in detecting 16S rRNA even at extremely low concentrations.

### Specificity analysis

For the specificity experiments, 16S rRNA targets from different samples were mixed with streptavidin-bound PQDs and biotinylated probes on a 96-well microplate reader plate. The microplate was arranged as follows: the first well served as a blank, containing 100 μl 1X PBS for reference. In the second well, 90 μl 1X PBS was added, followed by 10 μl PQDs. Subsequently, the third well-received 90 μl 1X PBS, along with the 10 μl streptavidin-conjugated PQDs. Careful pipette mixing was performed to ensure thorough mixing without touching the well walls. To assess specificity, different samples were then added to different wells to evaluate the specificity of the proposed sensing technique. After setting up the microplate, it was incubated to allow for interactions and reactions. Subsequently, fluorescence analysis was conducted using a multimode reader (Shandilya et al., 2022b). This analysis made it possible to test the specificity and binding abilities of the nanohybrid system with different PCR products. This gave us important information about its selectivity and how it could be used to find and analyze specific 16S rRNA.

## RESULTS AND DISCUSSION

In this study, we aimed to develop a fluorescence-based optical sensing system utilizing streptavidin-tethered PQD biotinylated oligonucleotide nanohybrid for the target-specific detection of 16S rRNA sequences in the CMB. To ensure the reliability of our experiments, we characterized the PQDs using the Malvern Zetasizer Nano ZS. The particle size analysis and zeta potential measurement were conducted on PQD using a Zeta Sizer instrument (Malvern Zetasizer Nano ZS). The Zeta Sizer utilizes dynamic light scattering (DLS) to determine the particle size distribution of the sample, and electrophoretic light scattering (ELS) to measure the zeta potential. The Z Average size representing the mean particle size based on the intensity-weighted size distribution of the PQD sample was determined to be 1880 d. nm. PQDs are known to possess nanoscale dimensions and an observed size of 1880 d. nm aligns with the expected size range for PQDs. The particle size distribution by intensity graph has been shown below represents the uniform size distribution of PQDs (Figure 3). The zeta potential of the PQDs was measured to be -6.57 mV. Zeta potential is a measure of the electric charge at the surface of the particles and provides information about the stability and dispersion behavior of the particles. The negative zeta potential indicates that the PQD particles have a net negative charge, which can be attributed to the presence of ionized functional groups on the surface of the quantum dots or the adsorption of ions from the surrounding medium also the negative charge is important for maintaining colloidal stability by providing electrostatic repulsion between particles, preventing agglomeration, and facilitating a stable dispersion. A high-resolution fluorescent microscope was used to examine PQD’s fluorescence. The PQDs exhibited high brightness, photostability, and uniformity in morphology, making them promising candidates for biosensing applications and ensured the acquisition of well-defined quantum dots with desirable properties for our nanohybrid construction.

**Figure 3:**
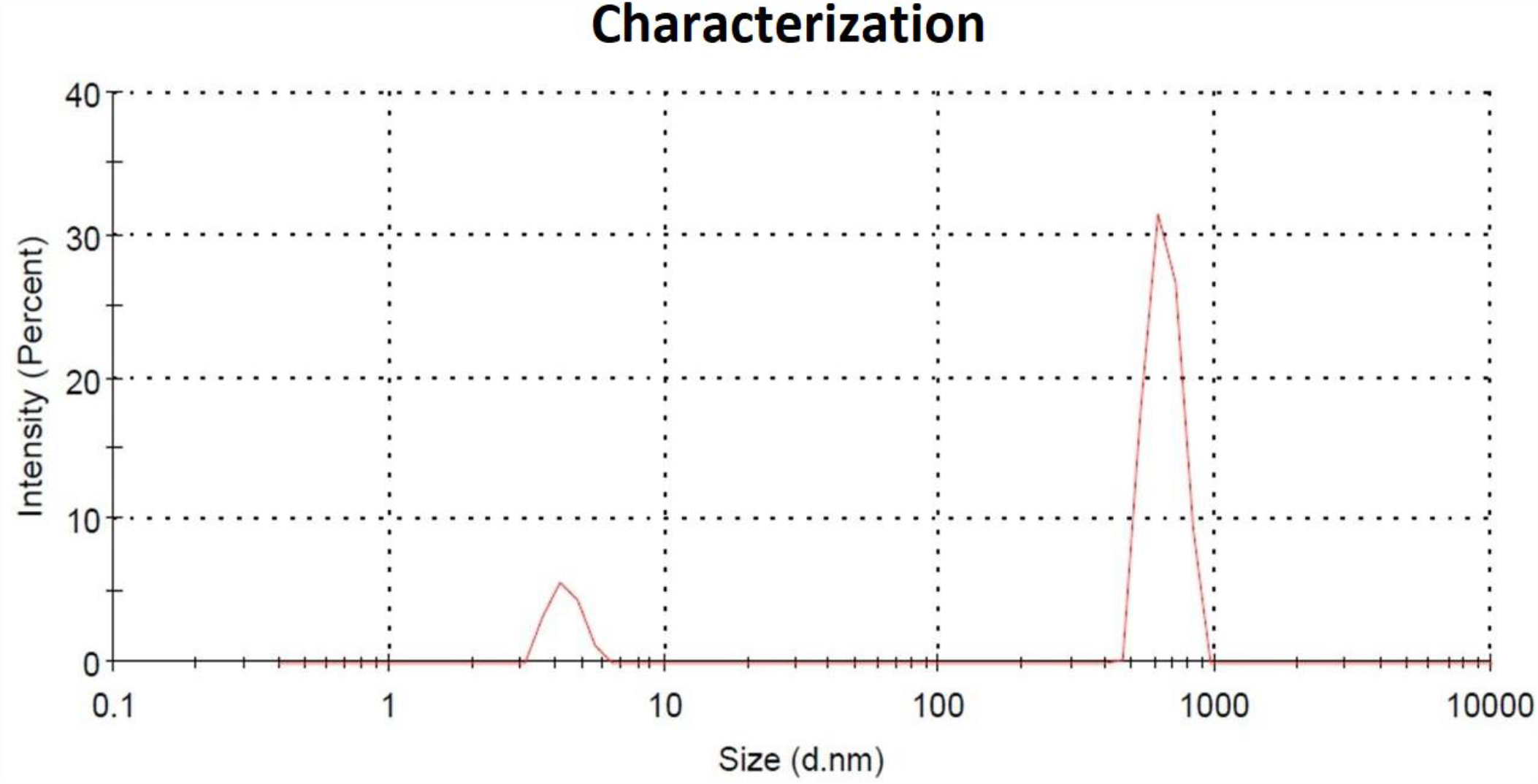
Figure showing the particle size distribution of PQDs characterized with Zeta-Sizer.

The 16S rRNA was isolated from the blood samples and further amplification was carried out through RT-PCR. The successful isolation and its subsequent amplification enable the further detection and analysis of CMB. Specific primers targeting long 16S rRNA fragments were used, resulting in adequately identifying different bacterial species within the sample. We created biotinylated oligonucleotide probes for our nanohybrid platform using the Pierce™ RNA 3’ End Biotinylation Kit. These probes are then directed to form a stable and targeted nanohybrid system by attaching to streptavidin-coated PQDs. Such interaction of the biotinylated probe ensures the target-specific binding to the target 16S rRNA sequences, providing a robust foundation for the subsequent detection experiments. We conducted high-resolution fluorescent microscopy and fluorometry analyses to evaluate the detection capability of our developed nanohybrid system.

Conjugation of PQD 450 and biotinylated probes resulted in strong and specific fluorescence signals localized to targeted 16S rRNA. The intercalating dye PI effectively inserts itself between the base pairs, emitting fluorescence when exposed to light of the appropriate wavelength, enabling visualization. Co-localization analysis revealed a significant overlap between PQD 450-biotinylated probe complexes and PI signals with the targeted 16S rRNA indicating potential interactions between them as described in Figure 4. These experiments confirmed the system’s effectiveness in detecting specific 16S rRNA sequences within the CMB samples. The visualization of specific 16S rRNA sequences in the fluorescence microscopy analysis demonstrated the nanohybrid system’s specificity and efficacy in identifying the target sequences.

**Figure 4:**
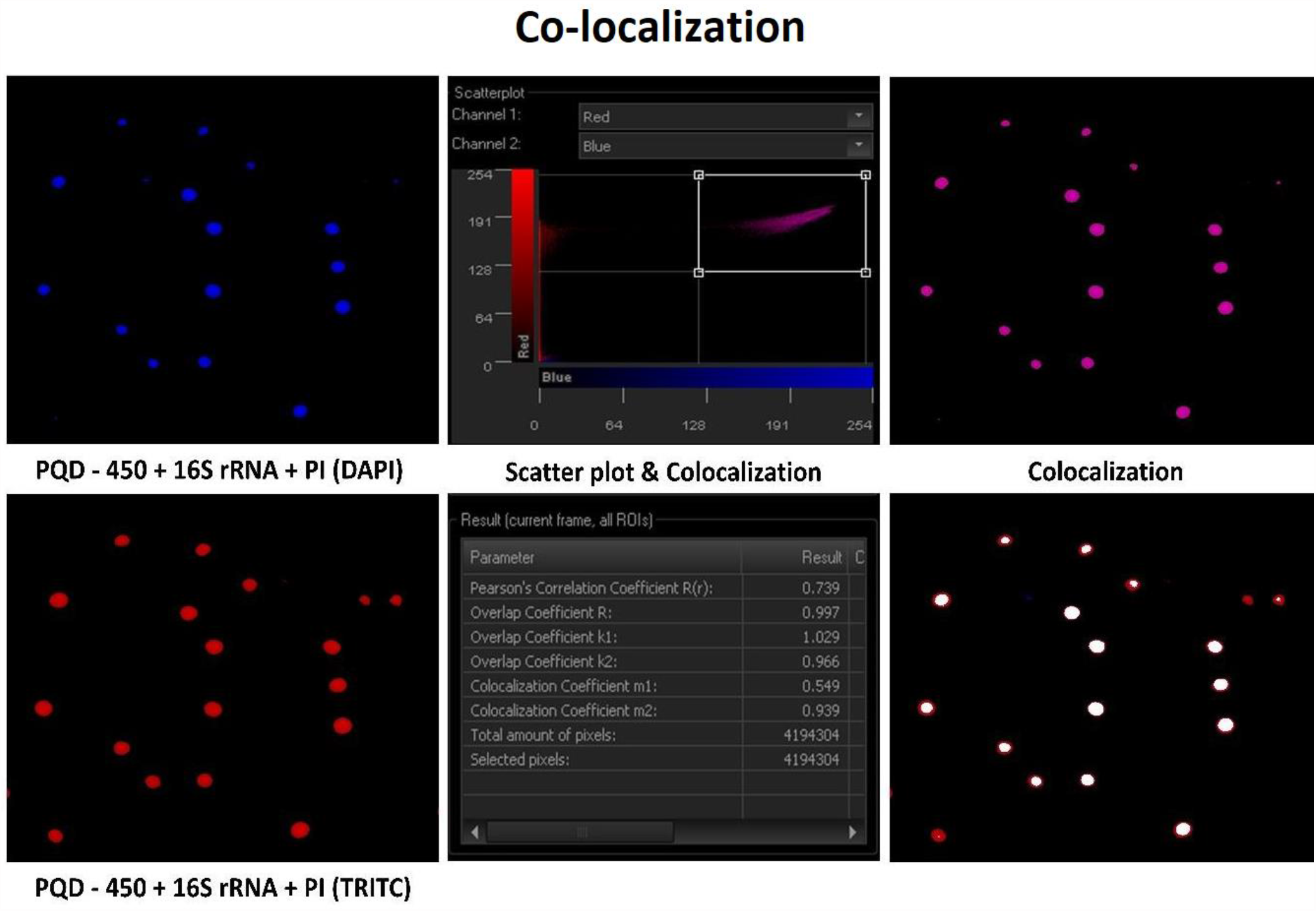
High-resolution microscopic analysis of the synthesized nanoanalytical framework.

A practical suitability assessment study was conducted using the Tecan Spark multimode reader to examine the efficiency of streptavidin-conjugated PQDs in the detection of 16S rRNA specifically. The results obtained from this experiment provided valuable insights into the applicability of our nanohybrid system, indicating its potential for future CMB research and analysis.

The applicability of the developed nanohybrid was described in Figure 5. The PQD shows exceptional fluorescence intensity (FI). The FI gradually declined with the conjugation of streptavidin to the surface of PQD. The attachment of streptavidin to the surface of PQD interferes with the fluorescence intensity of PQD. Quantum Dot possesses quantum confinement effect (QCE) within the 1-10 nm particle size. When the streptavidin-PQD conjugation takes place by COOH-NH2 coupling chemistry, the QCE was largely influenced by the change in particle size. This results in the alteration of the sp2 carbon domain core size. This will create energy gap alteration with the restriction of π-electron movement in the system. This phenomenon of changing fluorescence intensity is termed as “Blueshift”. When proteins are attached to the surface of PQDs, it affects the energy balance and generates Van der Waals forces. The added charge creates an electric field that blocks and compresses the effective volume of the PQDs. When biotinylated probes are attached, the compression increases and causes an enhancement in signal.

**Figure 5:**
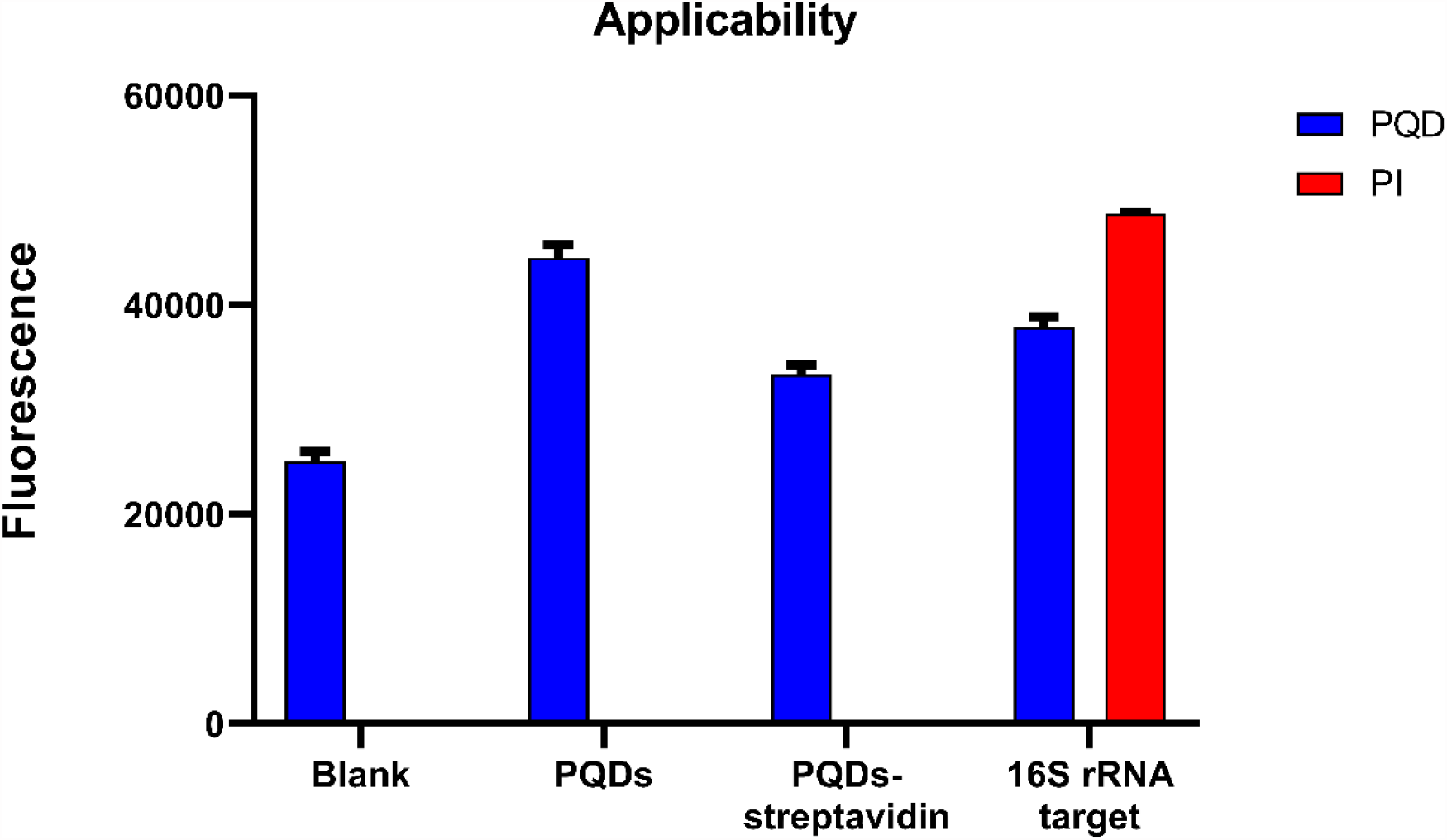
Graph showing applicability of the fabricated nanobiosensor.

The conjugated PQDs with streptavidin when complexed with a biotinylated oligonucleotide probe exhibited an increase in FI when specifically bound to targeted 16S rRNA in the presence of intercalating dye. Thus, the method is applicable for sensing the bacterial 16S rRNA. The sensitivity analysis we conducted aimed to estimate the nanohybrid system’s detection limit for 16S rRNA. The technique demonstrated exceptional sensitivity, detecting 16S rRNA even at deficient concentrations.This extraordinary sensitivity is encouraging for our nanohybrid system’s potential in clinical applications, where sensitive and precise detection was essential. The high sensitivity of the developed nanohybrid was found in the 1000ng of targeted biomolecule (16s rRNA) and exponentially decreases with decreasing the 16S rRNA concentration up to 1fg (Figure 6). The limit of detection (LOD) of this method for sensing bacterial 16S rRNA is sensitive up to 11.67 pg.

**Figure 6:**
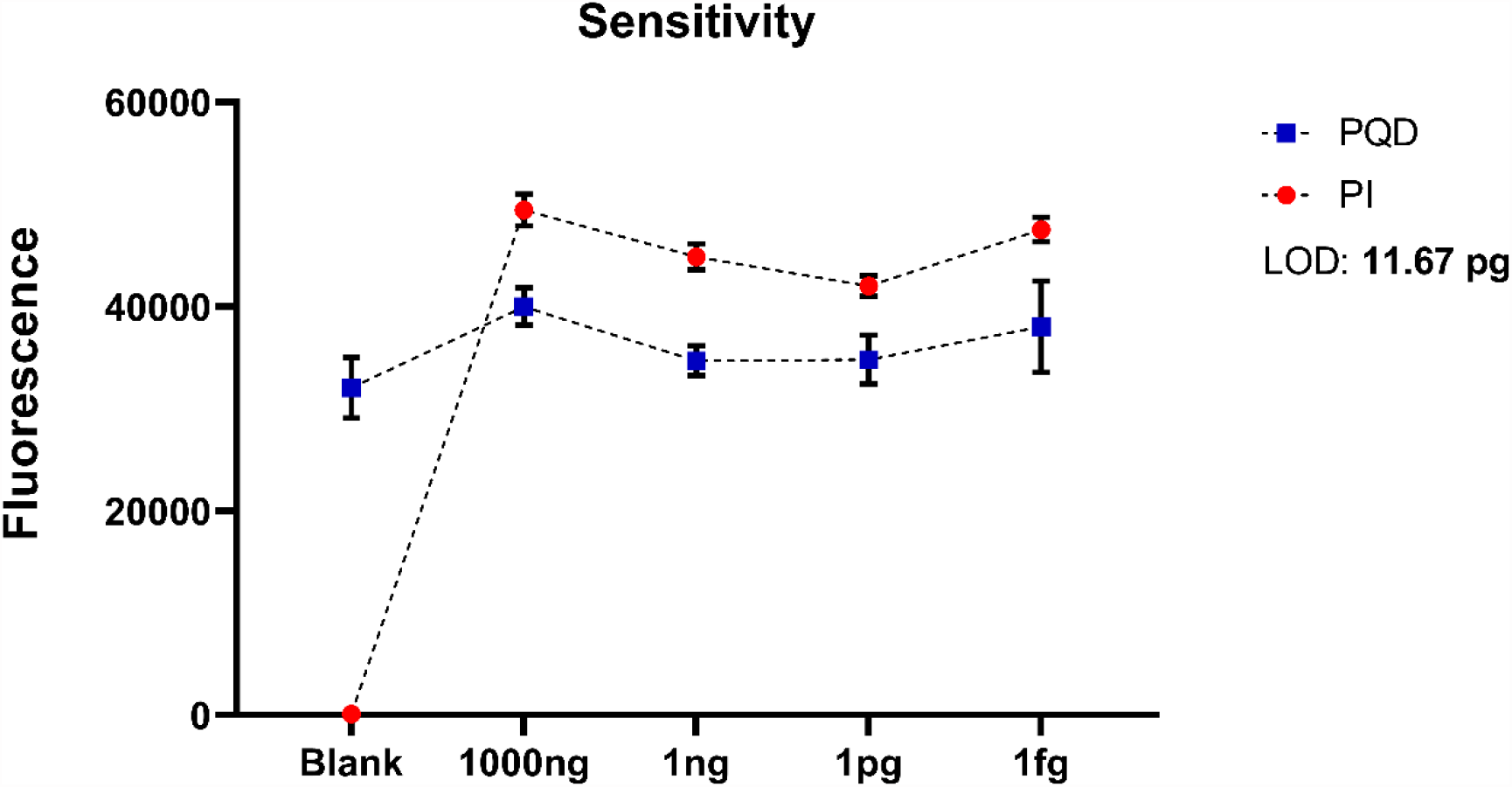
Graph showing sensitivity of the developed nanobiosensor.

Furthermore, a specificity study was performed to examine the nanohybrid system’s ability to differentiate different PCR products from various samples. The successful binding of the nanohybrid system with a specific 16S rRNA sequence amidst a complex biological matrix demonstrated its high selectivity and precision for target identification. In this method, two fragments were used for the specificity study with 16S rRNA. Human mitochondrial 16S rRNA (mt-16S rRNA) strands have similar sequences to bacterial 16S rRNA (Rehman et al., 2023). Figure 7 describes the nanohybrid structure that tends to bind bacterial 16S rRNA as compared to mt-16S rRNA. The size of the base pair of bacterial 16S rRNA is very much smaller as compared to mt-16S rRNA thus, having a high affinity towards the nanohybrid. In conclusion, our method is highly specific towards targeting bacterial 16S rRNA as a biomarker at most. This signifies the use of PQD-conjugated streptavidin-biotinylated oligonucleotide complex for the specific sensing of CMB in body fluids.

**Figure 7:**
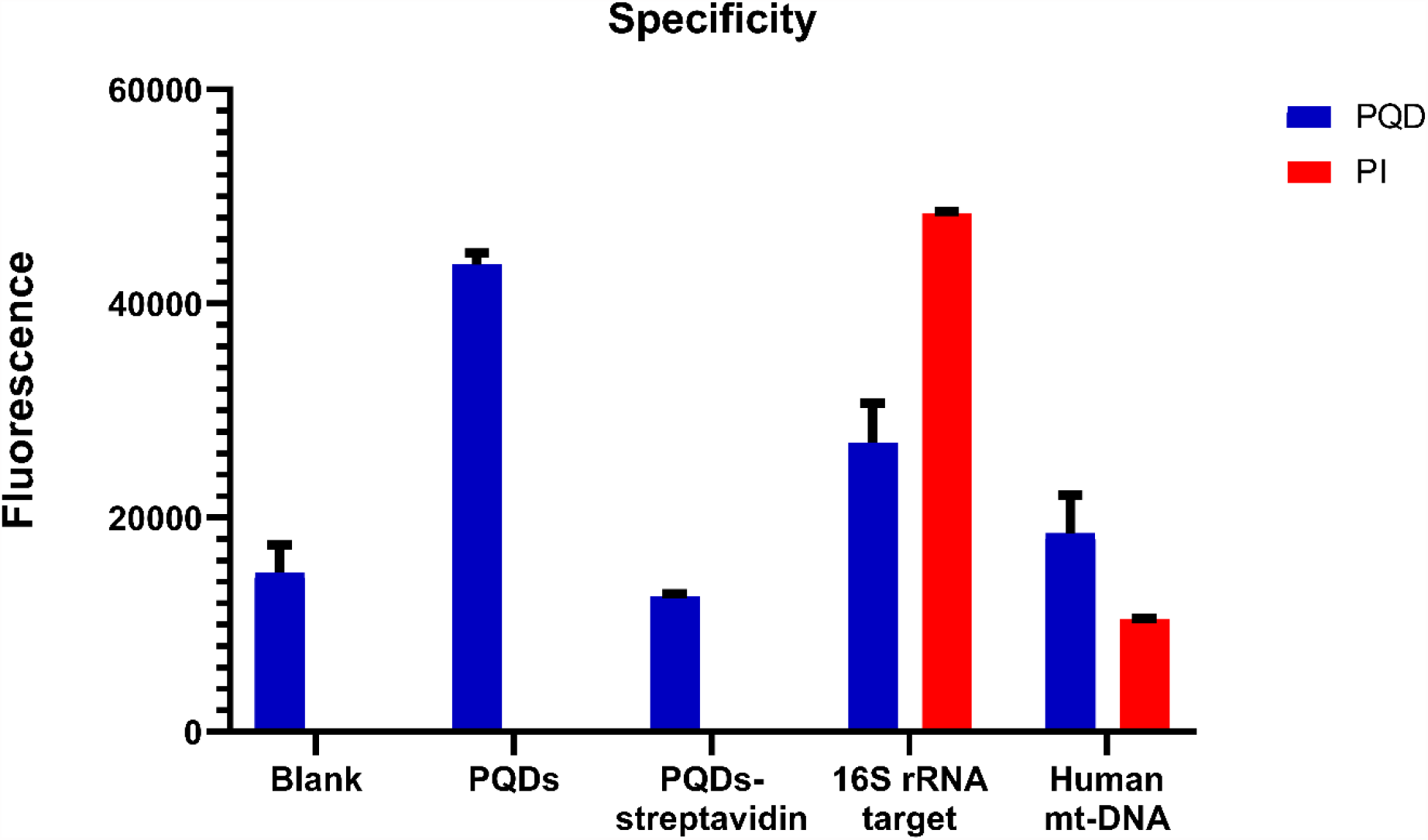
Graph showing specificity of the constructed nanobiosensor.

## CONCLUSION & TRANSLATIONAL PROSPECTS

Circulating cell-free microbiota has been the subject of significant attention as a possible biomarker for diagnosing non-communicable diseases, including CVD. Unfortunately, the current detection methodologies are hindered by time, funds, and expertise-related issues, which mostly prevent the therapeutic use of circulating microbiota. However, a solution may be on the horizon. Developments in point-of-care (POC) techniques, such as nanobiosensors, offer mobile on-site platforms, have a quick turnaround time, require minimal samples, and are cost-effective (Vermisoglou et al, 2020; Shandilya et al., 2021c). These characteristics could be a helpful solution to the current challenges in detection methodologies. The present study harnesses the remarkable photonic properties of PQDs to create a streamlined and fluorescence-based method for accurately identifying the CMB rapidly in clinical samples. Thorough characterization of the acquired PQDs and the utilization of specific PCR primers ensured the system’s reliability. The nanohybrid exhibited outstanding performance in sensing targeted 16S rRNA sequences. The results obtained in this study demonstrate the successful development of a robust fluorescence-based optical sensing system based on streptavidin-tethered PQD biotinylated oligonucleotide nanohybrid. This system shows great promise for the specific capturing of 16S rRNA sequences in the biosensing of CMB for CVD diagnosis. The exceptional sensitivity, specificity, and applicability make it an exciting tool for future clinical diagnostics and research involving CMB and also introduces new possibilities for comprehending the MB’s role in health and disease, presenting a potent tool for personalized medicine and healthcare research.

## Data Availability

All data produced in the present study are available upon reasonable request to the authors.

## Acknowledgements

The authors are thankful to the Indian Council of Medical Research (ICMR), Department of Health Research (DHR), Ministry of Health & Family Welfare (MoHFW), Government of India, New Delhi, for providing necessary financial support to the laboratory of Professor (Dr.) Pradyumna Kumar Mishra.

## Authors’ contribution

PKM devised the concept, developed the methodology, and supervised the experiments; NN, RD, and KZ performed the majority of experiments; VG collected and characterized samples; NN and VG designed the figures; NN, RT, and PKM performed the data analysis and interpretation; and NN, VG, and PKM drafted the original manuscript.

